# Analysis of 977 Long COVID Patients Reveals Prevalent Neuropathy and Association with Anti-Ganglioside Antibodies

**DOI:** 10.1101/2025.03.04.25323101

**Authors:** Cole Maguire, Kristina Kashyap, Elizabeth Williams, Rija Aziz, Maisey Schuler, Cheyenne Ahamed, Chumeng Wang, Aurelia Mena, Jeffrey Saniuk, Johanna Busch, Sara Austin, Mary Kelley, W. Michael Brode, Esther Melamed

## Abstract

**Background:** Long COVID (LC) is a novel condition that is characterized by persistent symptoms that last from months to years following a SARS-CoV-2 infection. While LC symptoms vary widely, neuropathy is one of the most prevalent symptoms and drastically affects patients’ quality of life. However, the underlying pathophysiology of LC neuropathy remains poorly understood. Here, we investigated the prevalence and potential mechanisms of LC neuropathy in the largest LC neuropathy cohort to date.

**Methods:** We conducted an observational study of 977 adults with LC at Dell Medical School. Participants underwent clinical assessments, skin punch biopsy, and comprehensive metabolic, endocrine and immunological profiling. A subset of patients received treatment with intravenous immunoglobulin (IVIG).

**Findings:** Neuropathic symptoms were reported by 55% (534/977) participants, with skin biopsy confirming small fiber neuropathy in 56.5% (48/85) cases, affecting both epidermal and autonomic nerve fibers. Common risk factors for neuropathy, including metabolic and endocrine disorders, did not fully explain neuropathic symptoms. While general immunological markers (lymphocyte, T cell, and B cell count and C reactive protein were unremarkable, unexpectedly, we detected anti-ganglioside antibodies (AGAs) in 25% of patients with LC neuropathy, a comparable rate to other AGA-associated neuropathies. Longitudinal testing revealed persistent AGA positivity, and multiple elevated AGAs in a subset of patients. In a pilot treatment cohort of eight patients, IVIG treatment resulted in improvement of patient reported neuropathic symptoms.

**Interpretation:** Our findings reveal a high prevalence of small fiber neuropathy in LC, with evidence suggesting an autoimmune mechanism involving AGAs in one in four LC neuropathy patients. The therapeutic response to IVIG further supports an autoimmune pathophysiology, suggesting potential benefits of immunomodulation in LC neuropathy patients.

## Introduction

Severe acute respiratory syndrome coronavirus 2 (SARS-CoV-2), a betacoronavirus identified in 2019, caused the emergence of Coronavirus Disease (COVID-19) and sparked an unprecedented global pandemic. Although typically associated with respiratory symptoms, it has become increasingly evident that COVID-19 is a complex multi-system disease^1–4^, commonly leading to neurological complications in acute infection^1,5–8^. Furthermore, extending beyond acute symptoms, at least 10% of COVID-19 patients go on to develop Long COVID (LC) as known as post-acute sequelae of COVID-19 (PASC), characterized by persistent symptoms for months to years following acute infection, with neurological symptoms as some of the most prevalent manifestations^9–15^.

Among the spectrum of LC neurological symptoms, neuropathy is very common, reported in 10-61% of LC patients^11,12,16–19^. LC patients’ neuropathic symptoms can include numbness, tingling, throbbing, and/or a piercing or burning pain, which can be associated with hyperalgesia (increased sensitivity to painful stimuli) and allodynia (pain from normally non-painful stimuli). Additionally, LC neuropathy symptoms often persist for months post-COVID, severely affecting patients’ quality of life^20,21^, and there are currently no effective therapies for this condition.

Despite its prevalence, the mechanisms underlying LC neuropathy remain poorly understood, and targeted treatments have yet to be systematically evaluated. Recent small studies suggest a heterogeneous pattern of peripheral nerve involvement, encompassing small fiber^19,22–28^, large fiber^19^, plexopathy^29^, or autonomic fiber damage^30^. Nerve injury has been hypothesized to arise due to *direct* viral neuroinvasion of peripheral nerves^31,32^ or *indirect* nerve injury resulting from neurovascular dysregulation^33^, persistent systemic inflammation, or autoimmunity^34–36^. However, the small sample sizes of prior studies have precluded definitive conclusions about the pathophysiological pathways associated with LC neuropathy.

In this study, we conduct a comprehensive longitudinal analysis of LC neuropathy in 977 patients with LC at a dedicated post-COVID clinic, to understand the prevalence and risk factors for LC neuropathy. Our results reveal that neuropathy is one of the most prevalent symptoms of LC, affecting 55% of patients. Importantly, we establish a novel association between LC neuropathy and anti-ganglioside antibodies, suggesting an autoimmune etiology. Furthermore, we report on benefits of intravenous immunoglobulin therapy (IVIG) in a small cohort of LC neuropathy patients. Together, these findings offer new insights into the epidemiology of LC neuropathy, providing evidence for an autoimmune mechanism highlighting immunomodulation as a promising treatment strategy.

## Methods

### Participant Recruitment and Consent

For retrospective electronic health record analysis, The University of Texas at Austin Institutional Review Board reviewed the study protocol and determined it met the criteria for exemption from IRB review under 45 CFR 46.104 (4) secondary research on data or specimens. For research blood samples, all participants gave written informed consent in adherence to all local and national regulations and the study was approved by the UT Austin Institutional Review Board (IRB ID: STUDY00002467).

### Retrospective Electronic Health Record Data Collection

Electronic health record data between June 2021 and January 2025 was obtained for LC patients of the UT Health Austin Post-COVID-19 Program Clinic in Austin, Texas. To qualify for care in the Post-COVID-19 Program, patients needed to be at least 12 weeks past their initial COVID-19 infection. While patients did not need to provide proof of a positive COVID-19 test, each person underwent screening by a specialized nurse. This screening verified that their medical history aligned with previous COVID-19 infection and that their ongoing symptoms were related to LC rather than other medical conditions.

As part of routine clinical onboarding, patients completed a physician-designed standard of care questionnaire via Research Electronic Data Capture (REDCap), including collection of demographics, history and severity of acute COVID-19 illness, and LC symptoms. Sex assigned at birth, race, and ethnicity were all patient reported. The list of LC symptoms and acute COVID-19 severity groups were adapted from the National Institute of Health (NIH), as previously described^11^.

Patient’s laboratory testing values (e.g. A1C, B vitamins, TSH, auto-antibody testing, etc) and medical history not captured in the REDCap as part of clinical onboarding were collected from their UT Health electronic health record by both manual review and the Dell Medical School Enterprise Data Intelligence Team. Laboratory results closest in time to the initial presentation to the Post-COVID Program clinic were used for each patient. For 82 LC neuropathy patients in the cohort, the results from a 3mm skin punch biopsy from both calf and thigh were also collected from their electronic health record. Briefly, the skin punch biopsies were sent to Therapath Neuropathology for small fiber neuropathy testing, which encompassed staining for PGP9.5 to quantify the density of small fiber nerves in both the epidermis and visible sweat gland(s), which were compared to sex and age-matched controls for expected density. For 3 additional LC neuropathy patients, previous clinical skin punch biopsy results from a different laboratory provider were used, but which lacked quantification of sweat gland nerve densities.

### Serum Processing, Cytokine Multiplexing, and Anti-Ganglioside Antibody Testing

Venous blood was collected in Greiner Bio-One Serum Clot Activating Tubes (Griener Bio-One 455010P) and was stored at room temperature vertically for at least 30 minutes before processing. The samples were then centrifuged at 2000 g and 4°C for 10 minutes, after which the serum was aliquoted and stored at -80°C.

Serum was sent to Eve Technologies to quantify 65 cytokines using Eve Technologies’ HD48A Cytokine Multiplexing assay. Anti-ganglioside antibody (AGA, cat no: 0051033) testing was conducted on serum by ARUP laboratories, using their clinical semi-quantitative enzyme-linked immunosorbent assay.

### Data availability

All data produced in the present study are available upon reasonable request to the authors.

## Results

In total, 977 patients with Long COVID (LC) were recruited from the Post-COVID-19 Program Clinic at UT Health Austin (Figure 1A). Of these 977 patients, 55% reported neuropathic symptoms (Figure 1B). The cohort was disproportionately female (66%), white (80.5%), non-Hispanic (79.9%), with an age range of 19-84 years old (median 46), reflective of the demographics of the Austin population (Table 1 and Figure 1C-D).

**Figure 1.**
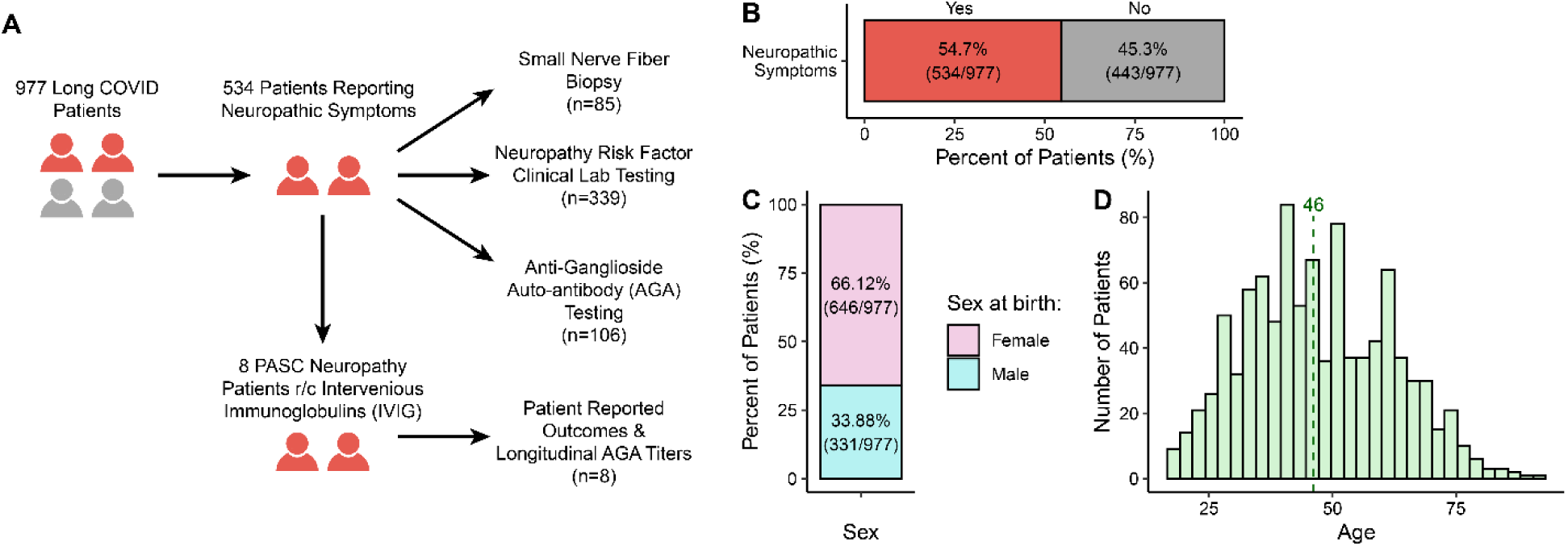
Demographics of LC and Neuropathy. **A)** Schematic of cohort and downstream analyses and sample sizes. **B)** Percentage of LC participants who reported neuropathic symptoms upon initial presentation. **C)** Sex ratio of the cohort. **D)** Age distribution of the cohort.

**Table 1.** Demographics of the Long COVID (LC) Cohort.

|  | Variable | LC Neuropathy<br>(n = 534) | LC No<br>Neuropathy<br>(n=443) | Total (n=977) |
| --- | --- | --- | --- | --- |
| **Sex at Birth** | Female | 71.5% (382/534) | 59.6% (264/443) | 66.1% (646/977) |
|  | Male | 28.5% (152/534) | 40.4% (179/443) | 33.9% (331/977) |
| **Age** | 18-29 | 11.8% (63/534) | 14.2% (63/443) | 12.9% (126/977) |
|  | 30-49 | 49.6% (265/534) | 40.6% (180/443) | 45.6% (445/977) |
|  | 50-70 | 35.0% (187/534) | 34.5% (153/443) | 34.8% (340/977) |
|  | >70 | 3.6% (19/534) | 10.6% (47/443) | 6.8% (66/977) |
| **Are you a person**<br>**of Hispanic, Latino,**<br>**or Spanish origin?** | No | 75.8% (405/534) | 84.9% (376/443) | 79.9% (781/977) |
|  | Yes | 21.9% (117/534) | 13.1% (58/443) | 17.9% (175/977) |
|  | Prefer not to answer | 2.3% (12/534) | 2.0% (9/443) | 2.2% (21/977) |
| **How do you define**<br>**your race?** | White, Caucasian | 79.0% (422/534) | 82.2% (364/443) | 80.5% (786/977) |
|  | Other | 6.2% (33/534) | 3.4% (15/443) | 4.9% (48/977) |
|  | Prefer not to answer | 5.4% (29/534) | 4.3% (19/443) | 4.9% (48/977) |
|  | Asian or Asian-American | 3.4% (18/534) | 4.7% (21/443) | 4.0% (39/977) |
|  | Black or African American | 3.4% (18/534) | 3.8% (17/443) | 3.6% (35/977) |
|  | American Indian or Alaska<br>Native | 1.7% (9/534) | 0.5% (2/443) | 1.1% (11/977) |
|  | Middle Eastern or North<br>African | 0.8% (4/534) | 1.1% (5/443) | 0.9% (9/977) |
|  | Native Hawaiian or Other<br>Pacific Islander | 0.2% (1/534) | 0% (0/443) | 0.1% (1/977) |
| **COVID-19 Severity** | No symptoms | 1.7% (9/524) | 2.2% (9/414) | 1.9% (18/938) |
|  | Mild | 41.4% (217/524) | 50.5% (209/414) | 45.4% (426/938) |
|  | Moderate | 47.3% (248/524) | 38.9% (161/414) | 43.6% (409/938) |
|  | Severe | 6.9% (36/524) | 6.5% (27/414) | 6.7% (63/938) |
|  | Critical | 2.7% (14/524) | 1.9% (8/414) | 2.4% (22/938) |
| **Comorbidities** | Asthma | 24.4% (129/529) | 18.1% (80/443) | 21.5% (209/972) |
|  | Autoimmune disorder | 14.9% (79/529) | 11.1% (49/443) | 13.2% (128/972) |
|  | Blood clots | 3.2% (17/529) | 1.1% (5/443) | 2.3% (22/972) |
|  | Blood disorder | 2.3% (12/529) | 1.8% (8/443) | 2.1% (20/972) |
|  | Cancer | 5.7% (30/529) | 5.9% (26/443) | 5.8% (56/972) |
|  | Chronic heart disease (not<br>hypertension) | 1.9% (10/529) | 0.7% (3/443) | 1.3% (13/972) |
|  | Chronic kidney disease | 1.1% (6/529) | 1.8% (8/443) | 1.4% (14/972) |
|  | Chronic liver disease | 1.5% (8/529) | 0.5% (2/443) | 1.0% (10/972) |
|  | Chronic lung disease<br>(COPD, or impaired lung<br>function not including<br>asthma) | 2.5% (13/529) | 2.0% (9/443) | 2.3% (22/972) |
|  | Diabetes | 8.1% (43/529) | 7% (31/443) | 7.6% (74/972) |
|  | Fibromyalgia or chronic fatigue | 11.0% (58/529) | 6.1% (27/443) | 8.7% (85/972) |
|  | HIV | 0.2% (1/529) | 0.5% (2/443) | 0.3% (3/972) |
|  | High blood pressure | 17.6% (93/529) | 17.8% (79/443) | 17.7% (172/972) |
|  | High cholesterol | 16.5% (87/529) | 17.2% (76/443) | 16.8% (163/972) |
|  | Immunodeficiency | 2.8% (15/529) | 1.4% (6/443) | 2.2% (21/972) |
|  | Mental health condition | 26.8% (142/529) | 23.9% (106/443) | 25.5% (248/972) |
|  | Migraines | 23.8% (126/529) | 14% (62/443) | 19.3% (188/972) |
|  | Stroke | 2.1% (11/529) | 0.5% (2/443) | 1.3% (13/972) |
|  | Substance use | 1.3% (7/529) | 2.5% (11/443) | 1.9% (18/972) |
| **Small Fiber Status** | SFN+ | 56.5% (48/85) |  |  |
|  | SFN- | 42.4% (36/85) |  |  |
|  | Indeterminate | 1.2% (1/85) |  |  |
| **Sweat Gland Nerve Density** | Reduced/Neuropathy | 29.3% (24/82) |  |  |
|  | Indeterminate | 37.8% (31/82) |  |  |
| **Epidermal Nerve Density** | Reduced/Neuropathy | 38.8% (33/85) |  |  |
|  | Indeterminate | 1.2% (1/85) |  |  |
| **Anti-ganglioside Antibodies** | Equivocal or Positive | 24.5% (26/106) | 0% (0/10) |  |
| **Elevated AGA by Ganglioside** | Asialo GM1 | 11.3% (12/106) | 0% (0/10) |  |
|  | GM1 | 6.6% (7/106) | 0% (0/10) |  |
|  | GM2 | 3.8% (4/106) | 0% (0/10) |  |
|  | GD1b | 3.8% (4/106) | 0% (0/10) |  |
|  | GD1a | 4.7% (5/106) | 0% (0/10) |  |
|  | GQ1b | 2.8% (3/106) | 0% (0/10) |  |

### Skin Punch Biopsies Demonstrate High Prevalence of Small Fiber Damage in Long COVID Neuropathy Patients

Out of 85 Long COVID neuropathy patients who underwent a skin punch biopsy, we observed a striking pattern of positive small fiber neuropathy (SFN). In this cohort, 56.5% (n=48/85) of patients demonstrated reduced small fiber nerve density in either epidermal and/or sweat gland fibers (Table 1 and Figure 2). Specifically, approximately half of patients (47.1%, n=24/51) had reduced sweat gland nerve density, when sweat gland(s) were present in the biopsy, and 38.8% (n=33/85) displayed diminished epidermal nerve density. Notably, 17.6% (n=9/51) of patients showed simultaneously diminished epidermal and sweat gland nerve density, underscoring the diffuse nature of peripheral nervous system damage in LC neuropathy. Further comparative analysis of biopsy sites revealed that distal biopsies (calf region) typically exhibited more pronounced small fiber loss compared to proximal sites (thigh region) (Figure 2, Supplemental Figure 2).

**Figure 2.**
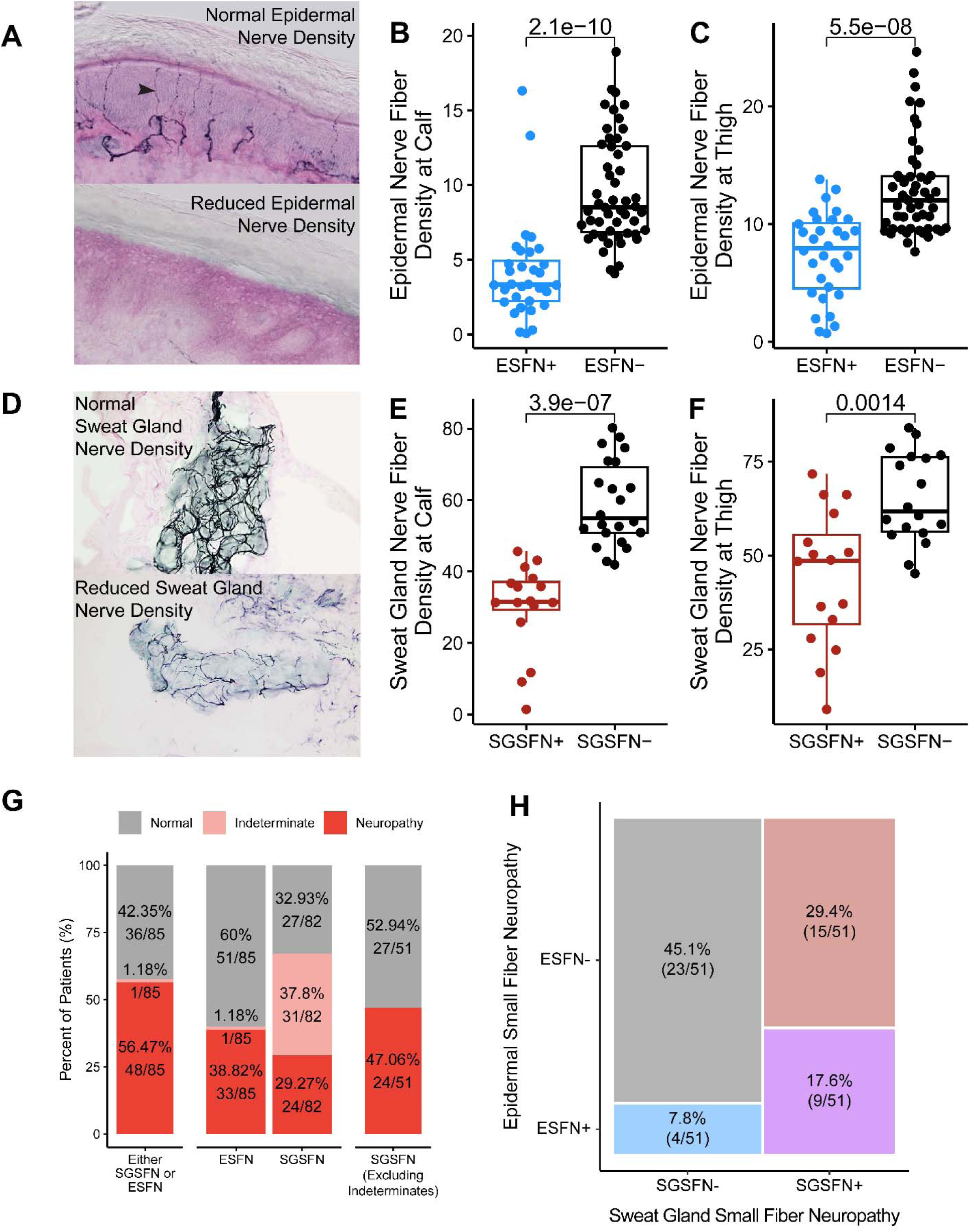
Small Fiber Neuropathy Affects both Autonomic and Sensory Peripheral Nerve Density in Long-COVID. **A)** Representative image of skin punch biopsy stained with PGP9.5 in a patient with normal epidermal nerve density and a patient with reduced density reflective of small fiber neuropathy. Representative images were provided by Therapath Neuropathology and used with permission. Epidermal nerve fiber density at the **B)** calf and **C)** thigh for patients that were epidermal small fiber neuropathy (ESFN) positive or negative. **D)** Representative image of skin punch biopsy stained with PGP9.5 in a patient with normal sweat gland density and a patient with reduced density reflective of small fiber neuropathy. Sweat gland nerve fiber density at the **E)** calf and **F)** thigh for patients that were sweat gland small fiber neuropathy (SGSFN) positive or negative. **G)** Percent of patients positive for either SGSFN or ESFN as well as percent of patients positive for specifically ESFN or SGSFN. Due to the high rate of indeterminates for SGSFN (often due to a lack of a sweat gland in the sample biopsied), the rate of SGSFN positivity was also calculated while excluding indeterminate cases. **H)** Mosaic plot demonstrating overlap of both SGSFN and ESFN in a portion of the cohort. Positivity was determined using Therapath’s sex and age adjusted cutoffs for nerve density in both epidermal and sweat glands, and patients were considered positive if either their calf or thigh sample was reduced.

### Long COVID Neuropathy Is Not Fully Attributed to Non-Autoimmune Causes

Common causes of peripheral neuropathy include diabetes, vitamin deficiencies, endocrine abnormalities, and toxic substances, among others^37^. To evaluate the extent to which these factors might account for the neuropathic symptoms in our patient cohort, we conducted a retrospective review of electronic health records for clinically available laboratory results. We found that the prevalence of diabetes in the neuropathy cohort (8.1%, 43/529) and elevated A1C (4.7%, 11/232, Figure 3A) were slightly below the rate of the general population (11.6% for diabetes^38^), suggesting that diabetes-associated neuropathy was not a primary cause in this cohort.

**Figure 3.**
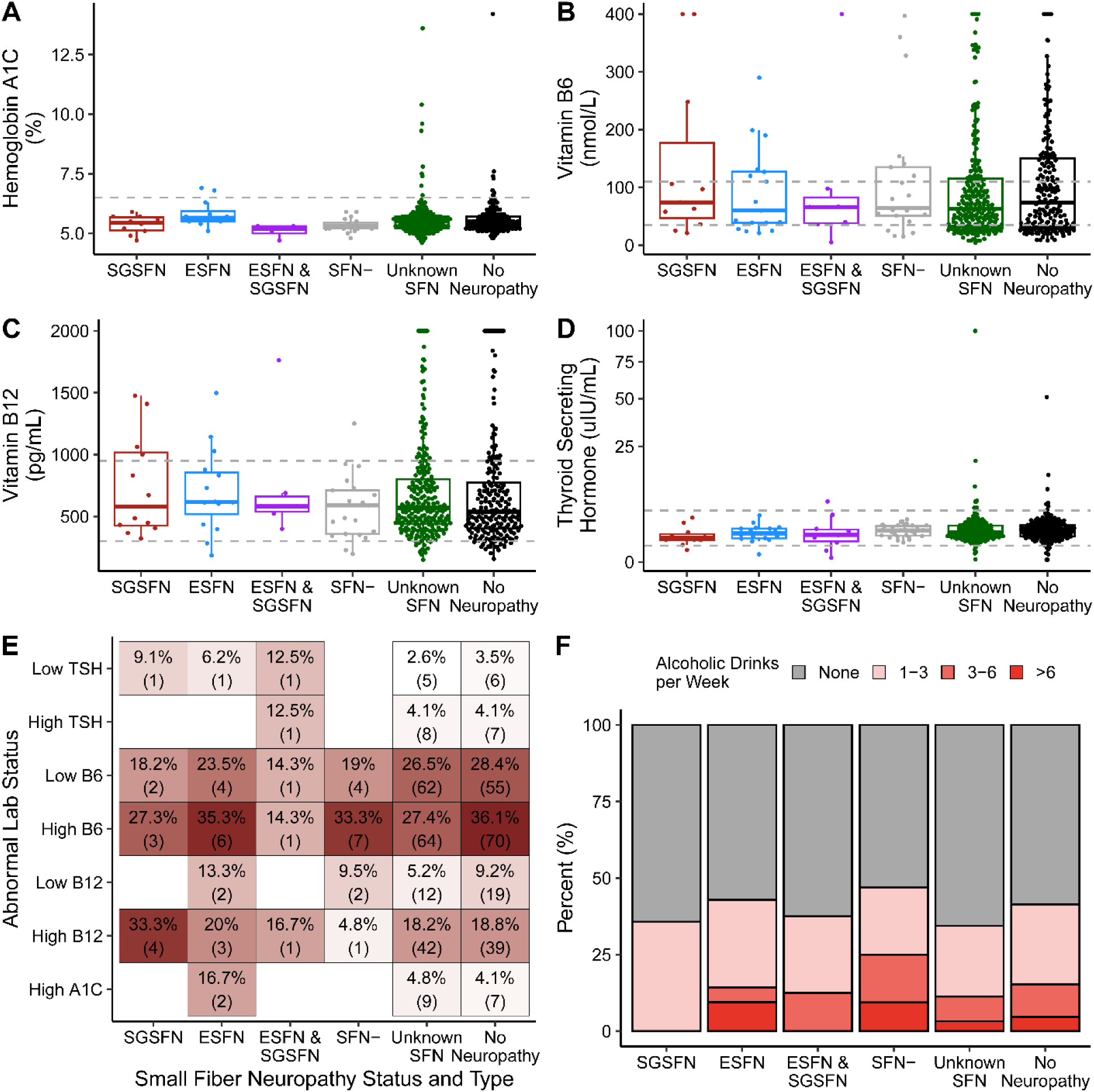
Diabetes, Vitamin Deficiency, Endocrine Abnormalities, and Alcohol Use Do Not Fully Explain LC Neuropathy. Boxplot of **A)** Hemoglobin A1C, **B)** Vitamin B6, **C)** Vitamin B12, and **D)** TSH by small fiber neuropathy status. Gray dotted lines denote clinical cut-offs for abnormal low and high status. **E)** Percentage of patients in each small fiber neuropathy group with laboratory values outside normal reference ranges for tests shown in panels A-D. **F)** Distribution of alcohol consumption patterns (by average weekly drink count) across small fiber neuropathy groups, showing percentage of patients in each consumption category.

Furthermore, we found that 27.9% (81/290) of patients had elevated B6 (>110 nmol/L, Figure 3B), 25.2% (73/290) has low B6 (<35 nmol/L, Figure 3B), 5.6% (16/285) had low B12 vitamins (<300 pg/mL, Figure 3C), 17.9% (51/285) had elevated B12 (>950 pg/mL, Figure 3C), and 6.6% had low (8/256, <0.5 uiU/mL, Figure 3D) or elevated (9/256, >5 uiU/mL, Figure 3D) thyroid stimulating hormone (TSH).

We also found that 3.6% (19/524) of all patients with neuropathic symptoms reported >6 alcoholic drinks per week, with 64.7% (319/524) reporting no alcohol consumption. In total, 38.3% of patients (130/339, only including patients who had at least one of A1C, B6, B12, and TSH evaluated) were negative for all evaluated risk factors. Interestingly, the rates of these neuropathic risk factors were comparable in LC patients with and without neuropathy, suggesting these factors may not play a critical role in the development of LC related neuropathy.

### Long COVID Neuropathy was Not Associated with Differences in Circulating Immune Cells and Cytokines

Given that common endocrine, metabolic or toxic causes did not fully explain the neuropathic symptoms in our cohort, we next investigated markers of inflammation and immune dysfunction, which can contribute to neuropathy. Of note, most LC neuropathy patients had normal absolute lymphocyte count (95.7%, 292/305, Figure 4A), absolute B cell counts (92.5%, 136/147, Figure 4B), absolute T cell count (93.9%, 139/148, Figure 4C), and C-Reactive protein (88.8%, 230/259, <1 mg/dL, Figure 4D). Interestingly, 11.7% (29/247) of LC neuropathy patients had low immunoglobulin levels, with 7.3% (18/247) having low IgM, 3.6% (9/247) low IgG, and 3.2% (8/247) low IgA (Figure 4E). Due to previous literature suggesting pro-longed inflammation in LC^39–44^, we also evaluated serum cytokine concentrations of 48 key cytokines, and observed no significance differences (Supplemental Figure 3). Although anti-nuclear antibodies, anti-TPO and anti-phospholipid antibodies were observed in ∼11-13% of participants, they were found at similar rates to the general population^45–47^ and are typically not associated with peripheral neuropathy (Figure 4F). Thus, overall common immunological measures were unremarkable in patients with LC neuropathy.

**Figure 4.**
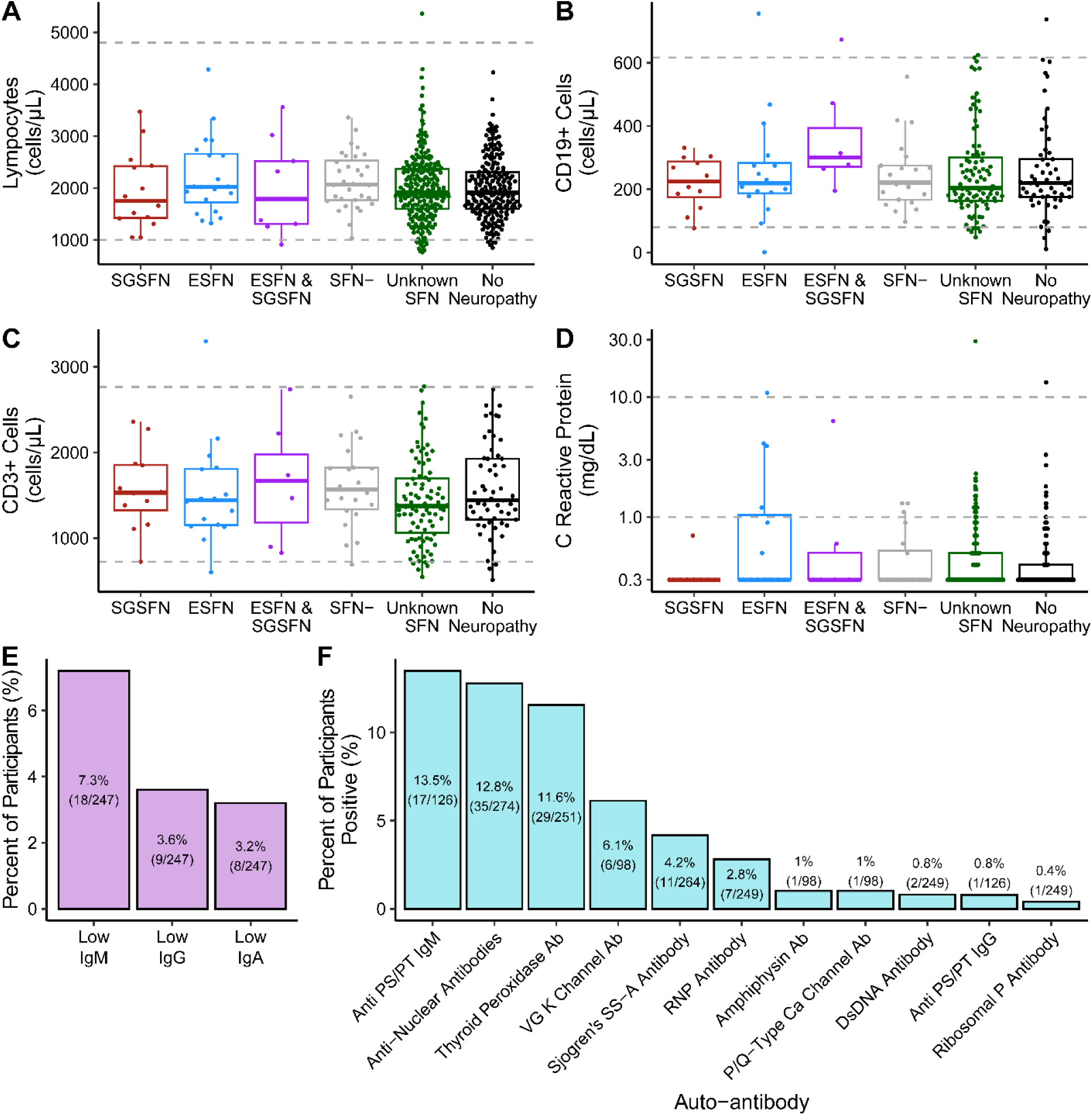
No distinctive general immunological markers identified in LC Neuropathy. **A)** Absolute lymphocyte count, **B)** CD19+ B-cell count, **C)** CD3+ T-cell count, and **D)** CRP by reported LC neuropathy symptoms and skin punch biopsy findings where available. **E)** Percent of LC neuropathy patients with low immunoglobulins by Ig isotype. **F)** Percent of LC neuropathy patients with available clinical autoantibody data.

### High Prevalence of Anti-Ganglioside Antibodies in LC Patients with Neuropathic Symptoms

Given the established role of autoimmunity in peripheral neuropathies^48^ and the evidence that acute COVID-19 infection can induce autoimmunity^49,50^, we next evaluated for known auto-antibodies associated with post-infectious neuropathies. Specifically, we focused on anti-ganglioside antibodies (AGAs, Figure 5A), which are commonly observed in Guillan Barré Syndrome (GBS), a post-infection-associated peripheral autoimmune neuropathy^51,52^ and in multifocal motor neuropathies (MMN), a progressive demyelinating immune-mediated neuropathy.

**Figure 5.**
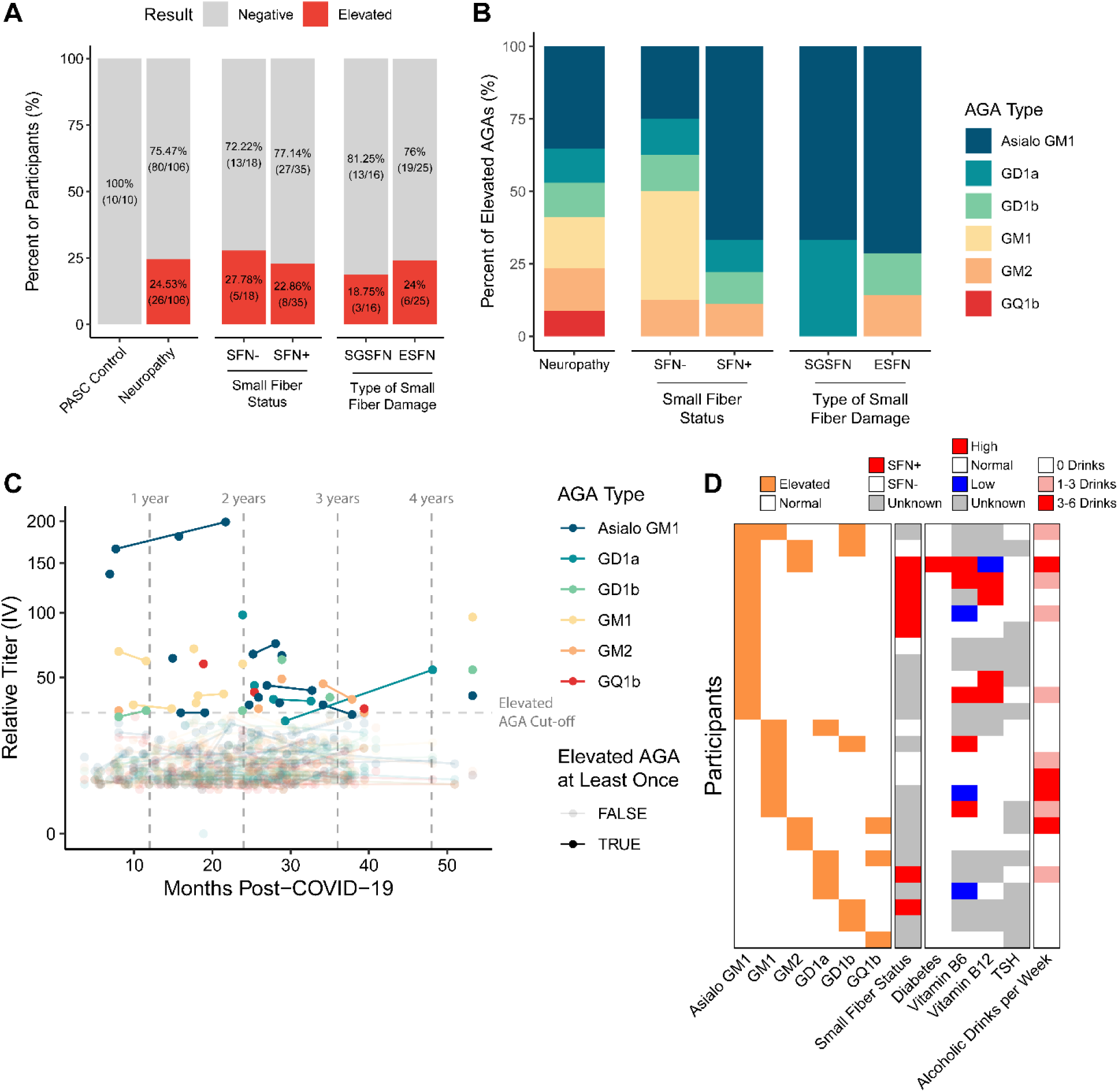
Anti-Ganglioside Antibodies are Found in One in Four LC Neuropathy Patients. **A)** Percent of LC Neuropathy patients with elevated AGAs, with subanalyses by small fiber neuropathy (SFN) status (confirmed by skin punch biopsy) and SFN subtype. **B)** Distribution of elevated AGAs by ganglioside antigen, stratified by SFN status and subtype. **C)** Longitudinal analysis of AGA titers from initial COVID-19 infection. **D)** Heatmap showing overlap of neuropathy risk factors with elevated AGAs, suggesting possible synergistic effects. Each row denotes an individual participant.

In total, we evaluated AGAs in 169 serum samples from 116 LC patients with (n=106) and without neuropathy (n=10) as part of active clinical care or in longitudinal bio-banked samples. Notably, we found that approximately one in four LC neuropathy patients in our cohort (24.5%, n=26/106) had elevated AGAs (>29 IV) compared to none of the control LC patients without neuropathy (0%, 0/10) (Figure 5A). These elevated AGA results ranged across a spectrum of values, with 42% (11/26) measuring above 50 IV and the remaining 58% (15/26) falling within the 30-50 IV range. The presence of AGAs was independent of small fiber neuropathy status and was found in both SFN+ and SFN-patients (Figure 5A). Within the SFN+ patients, we did not find an association between AGA positivity and specific type of small nerve fiber involvement (i.e. epidermal or sweat gland) (Figure 5A).

Interestingly, we found distinct patterns of AGAs between SFN+ and SFN-neuropathy patients (Figure 5B). For example, SFN positivity was associated with a higher percentage of Asialo GM1 AGAs, whereas SFN negative patients had higher GM1 and GM2 AGA positivity.

Using bio-banked serum, we then repeated AGA testing on longitudinal patient samples who were found to be AGA positive (Figure 5C). Interestingly, after patients’ initial positive AGA result, we observed that AGA titers always persisted in subsequent samples, even extending up to ∼1.5 years apart. Thus, AGA titers were consistent overtime, highlighting one possible mechanism for the persistence of neuropathic symptoms. These findings suggest that AGAs are widely prevalent in Long-COVID neuropathy and are not just a transient phenomenon. Thus, the persistence of AGAs could contribute to Long-COVID neuropathy and further explain why neuropathic symptoms persist in this patient population.

Finally, we observed that other risk factors for neuropathy such as B vitamin abnormalities, were frequently observed in AGA+ patients, suggesting potentially synergistic causes of neuropathy in LC.

### Non-neuropathic LC symptoms are not associated with SFN or AGA positivity

Since LC is often characterized by multiple and varied symptomatic sequalae^15,49^, we next examined the association of both SFN and AGA positivity with other typical LC associated symptoms (Supplementary Figure 4). This analysis did not yield any significant associations between non-neuropathic LC symptoms and either SFN or AGA status, however associations of other symptoms with AGAs, may be dependent on the ganglioside for which there is autoreactivity.

### Intravenous immunoglobulin (IVIG) alleviates LC Neuropathy Symptoms

IVIG is an intravenous infusion of isolated immunoglobulins combined from hundreds of healthy individuals and is routinely used as an immunotherapy for autoimmune diseases^53^. Given the common use of IVIG for AGA associated neuropathies, such as GBS, we next investigated whether IVIG could alleviate LC neuropathy symptoms. Eight patients in our LC neuropathy cohort received IVIG therapy (1.2 g/kg monthly for 3-12 months). These patients qualified for IVIG therapy if they had an underlying immunological condition for which IVIG is FDA approved.

All patients reported at least some improvement of neuropathic symptoms post-IVIG, with 37.5% (3/8) reporting good improvement (4-point improvement on a 1 to 5 scale), and 62.5% (5/8) of patients reporting some improvement (3-point improvement on a 1 to 5 scale) (Figure 6A). Interestingly, 25% (2/8) patients were positive for AGAs, with both patients’ titers persisting post IVIG treatment (Figure 6B). Thus, these results suggest that IVIG may have a therapeutic benefit that is independent of affecting the AGA titer and may act via other known immune mechanisms^53^.

**Figure 6.**
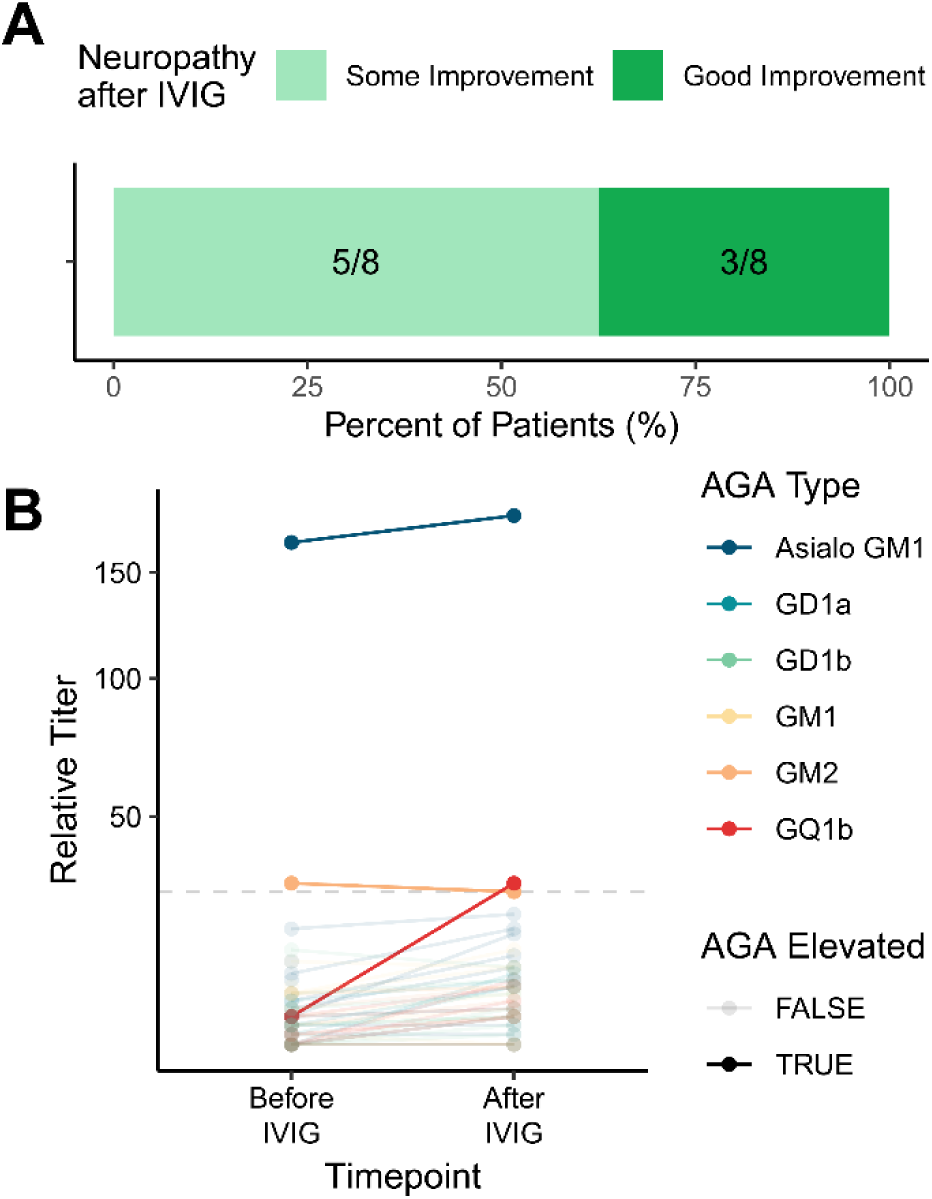
IVIG Alleviates Neuropathic Symptoms in a Small Cohort of LC Neuropathy Patients. **A)** Percent of participants reporting some improvement (3-point improvement on a 1 to 5 scale) or good improvement (4-point improvement on a 1 to 5 scale) to their neuropathic symptoms following IVIG. **B)** AGA titers before and after IVIG administration revealed constant titers. Of note, one participant positive for GM2 also developed an elevated GQ1b overtime.

## Discussion

In this study, we examined a cohort of 977 Long COVID (LC) patients at a dedicated Post-COVID-19 Program Clinic and found that 55% reported neuropathy symptoms. Detailed evaluation of skin punch biopsies in 85 LC neuropathy patients demonstrated that 38.8% had epidermal and 47% displayed sweat gland fiber abnormalities, demonstrating that PASC neuropathy is frequently associated with direct damage to both somatic and autonomic small fiber nerves. A key finding of our study was the detection of elevated anti-ganglioside antibodies (AGAs) in approximately one in four LC neuropathy patients and none of the controls, providing strong evidence for an autoimmune mechanism in LC neuropathy. Among the six detected AGAs (GM1, Asialo GM1, GM2, GD1a, GD1b, GQ1b), GM1 and Asialo GM1 were predominant, while GD1 Ab’s demonstrated a specific association with sweat gland small fiber neuropathy. This represents a striking enrichment of autoimmune-mediated neuropathy in this patient population, exceeding the expected rate of AGA’s (∼1%-12%) reported in the general population^54,55^. The persistence of AGAs over time, even up to 1.5 years post-infection, underscores their potential role in chronic symptomatology of LC neuropathy. Additionally, all eight LC neuropathy patients treated with IVIG reported marked symptomatic improvement, suggesting that IVIG or other immunomodulatory therapies could offer promising treatment options for this population. Overall, our manuscript is novel in establishing a link between autoimmunity and LC neuropathy, and providing direct evidence for benefits of targeted immunomodulation with IVIG.

### Box: Background on Peripheral Neuropathy

Peripheral neuropathies can result in a variety of symptoms affecting sensory, motor, and autonomic function, depending on the involved nerve fibers. Peripheral nerve fibers are broadly categorized as small or large. Small fibers are typically responsible for pain, temperature, and autonomic function (e.g. sweating). Conversely, large fibers control sensation of light touch, vibration, joint position, and muscle movements. Large fiber neuropathies are typically confirmed with nerve conduction studies, while small fiber neuropathies require a skin punch biopsy. Neuropathic symptoms can result from either small or large fiber neuropathies, or both, and thus, require a detailed assessment to evaluate the potential causes.

The etiologies of peripheral neuropathy encompass over 100 different conditions. Diabetes is the most common cause, found in 18-49% of cases, followed by idiopathic/cryptogenic causes in 12-49%^56–61^. Other major causes include high alcohol consumption (1-19%), inflammatory conditions (1-16%), genetic disorders (1-14%), toxic/medication-induced (3-14%), and nutritional deficiencies (1-9%), particularly vitamin B6 and B12 deficiencies or excess. Less commonly, neuropathies are caused by paraproteinemias (1-9%), infections (up to 12%), connective tissue disorders/vasculitis (1-5%), and renal failure (1-4%)^62–69^. Rare causes include malignancy (1-13%), hypothyroidism (1-4%), sarcoidosis (0-1%), and liver disease (0-1%)^62–69^.

Antibodies against gangliosides, a class of glycosylated lipids enriched in the cell membranes of neurons and myelinating cells, have been described in a wide spectrum of neurological conditions in the peripheral and central nervous system (PNS and CNS)^70^. In the PNS, AGAs have distinct effects based on the nerve fibers they target. They are primarily associated with demyelinating conditions like GBS and MMN, which affect large motor nerves, leading to motor dysfunction^71^. AGAs can also target unmyelinated small fibers, causing SFN or sensory ataxic neuropathy^72,73^. In CNS-related disorders, such as Alzheimer’s disease and amyotrophic lateral sclerosis (ALS), AGAs are detected, though at lower rates than in peripheral neuropathies and are not thought to be reliable diagnostic markers in these conditions^74,75^.

As gangliosides are concentrated in the nodes of Ranvier and paranodal regions, the binding of AGAs is thought to disrupt axonal conduction through i) interfering with protein complexes such as critical ion channels^76^, or ii) result in complement deposition leading to axonal degeneration^77^. Additionally, endothelial cells expressing gangliosides can be targeted, resulting in vascular inflammation and compromised blood-nerve barrier integrity, exacerbating the immune-mediated injury and evoking further immune infiltration^78^.

While their pathological effects are established, the triggers for AGA production across diseases remain unclear. One potential mechanism is molecular mimicry, where pathogens mimic host molecular structures capable of triggering cross-reactive immune responses^79^. Molecular mimicry is well-documented in GBS, particularly for the bacteria *Campylobacter jejuni* and *Haemophilus influenzae* or chronic infecting herpes viruses such as human cytomegalovirus (CMV)^80–82^. Interestingly, recent evidence has demonstrated that COVID-19 leads to reactivation of chronic viral infections^83^, with this phenomenon linked to the development of LC^43,84^. Thus, molecular mimicry might serve as one potential mechanism for the formation of AGAs in LC neuropathy.

Of note, in the peripheral nervous system, AGAs are primarily associated with GBS, although the rate of AGA positivity depends on the specific subtype of GBS. For example, in the most common variant, acute inflammatory demyelinating polyneuropathy (AIDP), AGAs are only found in 6% of patients^85^, where as in acute motor axonal neuropathy and Miller-Fisher subtypes, AGAs are found in 40-80% of patients^85,86^. Although AGA positivity varies widely in post-viral neuropathies. AGAs serve as a well-accepted clinical biomarker that has been shown to contribute to disease pathology^87–90^. Our finding of elevated AGAs in 25% LC neuropathy patients is comparable to other AGA-associated neuropathies and suggests that AGAs may be a useful clinical diagnostic marker to identify LC patients who might respond to immunotherapies, such as IVIG, commonly used for GBS.

Beyond total AGA positivity rates, AGA subtypes have been associated with specific neuropathies. For example, anti-GQ1b AGAs have been linked to Miller-Fisher syndrome (rare variant of GBS)^91^, while GM1 and GM2 have been associated with chronic neuropathies, such as MMN^61^. In our cohort, the most commonly detected AGAs were Asialo GM1 (34%) and GM1 (18%), although elevated antibody titers were detected for all six AGAs. Interestingly, the ratio of Asialo GM1 to GM1 differed between SFN+ and SFN-patients, with Asialo GM1 more commonly found in the SFN+ patients.

Notably, while elevated, the AGA titers in our cohort, while still above the clinical threshold, were lower compared to other AGA-associated neuropathies like GBS, which notably have transiently high AGA titer^92^. Interestingly, titer levels of autoantibodies in neuroimmune conditions frequently affect types of symptoms and their clinical presentation. For example, autoantibodies against glutamic acid decarboxylase 65 (GAD65) at different titers may be associated with either autoimmune encephalitis, Stiff person syndrome, or type 1 diabetes^93^. Thus, differences in both AGA types as well as titer levels between AGA-associated neuropathies, may explain variations in severity and presentation of neuropathic symptoms. As a result of the variation in titers, the clinical thresholds used to determine AGA positivity may need to be modified as they were originally designed for GBS, a more severe neuropathy. Importantly, as previous studies have extensively demonstrated the direct pathogenicity of AGAs in causing nerve damage ^87–90^, our finding of AGA antibodies, even at lower titers, in LC neuropathy suggests that autoantibody-mediated nerve damage could be a key mechanism in LC neuropathy. Future studies in patients and animal models could confirm the causative role of AGAs in LC neuropathy.

Among the SFN+ participants, the distal samples (calf) displayed more pronounced loss of nerve fibers than samples collected more proximally (thigh). This pattern of findings on skin punch biopsies is consistent with a length dependent neuropathy. Furthermore, the distal samples more frequently captured sweat glands, enabling more consistent measurements of sweat gland nerve density compared to biopsies from the thigh. These findings suggest that skin punch biopsy in LC may be more informative and provide more specific physiological insights from distal rather than proximal sites.

Additionally, we observed elevated AGAs in 22.9% of SFN+ patients. This finding is intriguing as AGAs have traditionally been primarily associated with large fiber neuropathies, including demyelinating conditions and axonal neuropathies^51,52,71^. Our observation of AGAs in SFN+ raises a novel possibility that AGAs may play a broader pathophysiological role in both large and small fiber neuropathies. Additionally, future studies should evaluate additional autoantibodies to small and large nerve fibers beyond AGAs, which could reveal a larger contribution of autoimmunity in LC neuropathy and may explain the diverse symptomatology of LC neuropathic symptoms.

While metabolic abnormalities, such as low or elevated vitamin B6 levels, low or elevated B12 levels, elevated TSH, and diabetes, were frequently observed in our study population, they occurred at comparable rates in LC participants with or without neuropathy. Thus, metabolic factors alone may not explain all cases of LC neuropathy. Furthermore, these metabolic derangements were frequently observed concomitantly with AGAs, indicating that multiple mechanisms may potentially contribute to LC neuropathy, with both metabolic and autoimmune mechanisms potentially acting synergistically.

Notably, in a small cohort of eight LC neuropathy patients treated with IVIG, all patients reported improvement in neuropathic symptoms following treatment. Two of these patients were found to have AGAs prior to IVIG, and their AGA titer did not decrease following therapy. Thus, these results suggest that IVIG’s therapeutic benefits in LC neuropathy may derive from its known immunomodulatory effects such as neutralizing cytokines and autoantibodies, saturating Fc receptors, blocking complement activation, and regulating T and B cell inflammatory responses (reviewed in ^53^) rather than reducing autoantibody titers. In addition, IVIG has established efficacy in both AGA-associated neuropathies, such as AIDP, as well as non-AGA neuropathies, such as chronic inflammatory demyelinating polyneuropathy. Small case series have demonstrated the efficacy of IVIG in LC^94^, and there are currently large ongoing clinical trials for both IVIG and subcutaneous immunoglobulin in LC-associated postural orthostatic tachycardia syndrome (POTS) (NCT06305793 and NCT06524739, respectively). Our study provides the first evidence for IVIG efficacy specifically in LC neuropathy, supporting the rationale for larger clinical trials of immunoglobulin therapy in the LC neuropathy population.

Our study had several limitations. First, our reported rate of elevated AGAs in LC neuropathy may be an underestimate of the actual AGA rate in the LC patient population, due to the cutoffs for positivity on the clinical ELISA primarily informed by AIDP studies, a large-fiber neuropathy, which may potentially result in higher AGA titers than in LC neuropathy. Second, the currently available AGA clinical test is limited to only six ganglioside variants, thus raising the possibility that some patients may mount titers against other neuronal-associated gangliosides not available for clinical testing. Third, although we observed decreases in the sweat gland nerve density suggestive of dysautonomia, we did not formally evaluate for autonomic dysfunction in all patients, which would be important for future studies of this patient population. Fourth, our cohort of neuropathy patients that received IVIG was small (n=8) and was not a randomized control trial but serves as preliminary evidence for a randomized controlled trial in the future. Lastly, although reflective of the clinic population, our cohort was largely Caucasian (80.5%), female (66.1%), and non-Hispanic (79.9%), making it important to continue to evaluate more diverse patient populations in future studies of LC neuropathy.

In conclusion, our study provides the most comprehensive analysis of LC neuropathy patients to date, incorporating medical history data and standardized laboratory assessments as well as skin punch biopsies that enable clear differentiation between various neuropathy etiologies. Our longitudinal study design provides key temporal data demonstrating the persistence of AGAs in LC neuropathy patients, offering important insights into the chronic nature of this condition. In addition, we employed a clinically validated bioassay for AGA detection, making our findings immediately applicable and readily translatable in clinical practice. Altogether, our study provides the first evidence for autoimmune pathophysiology in LC neuropathy and demonstrates the therapeutic potential for immunomodulating therapies in the LC patient population.

## Data Availability

All data produced in the present study are available upon reasonable request to the authors.

## Acknowledgements

Firstly, we would like to thank and acknowledge the Long COVID patients at the UT Health Austin Dell Medical School Post-COVID program clinic, and participants who provided research blood samples, without whom this work would not be possible. We also appreciate technical support from April Demeo and Monica Deza at Clinical Pathology Labs. Additionally, we are grateful for the administrative support from Dell Medical School Neurology Department, particularly Sage Shaw.

This work was supported by The University of Texas at Austin Graduate School Continuing Fellowship (C.M.), philanthropic funds from Mr. Tom Bonney (E.M), NIAAA K08AA027837-05 (E.M.), The University of Texas at Austin Research and Creative Grant Award (E.M.), and institutional Dell Medical School Startup funding (E.M.). Funding sources did not have a direct role in design, analysis, or approval of this manuscript.

## Competing Interests

EM is a principal investigator on a subcutaneous immunoglobulin clinical trial (CSL-Behring) for post-COVID Postural Orthostatic Tachycardia Syndrome. WMB and JB are co-investigators on the same study. The other authors declare no competing interests.

## Notes

### Summary of Updates

Figure 2G was updated to clarify the rate of indeterminate results from skin punch biopsy; minor changes to the text to clarify existing statements; grammatical errors were corrected.

