## Supplement for "Analysis of 977 Long COVID Patients Reveals Prevalent Neuropathy and Association with Anti-Ganglioside Antibodies"

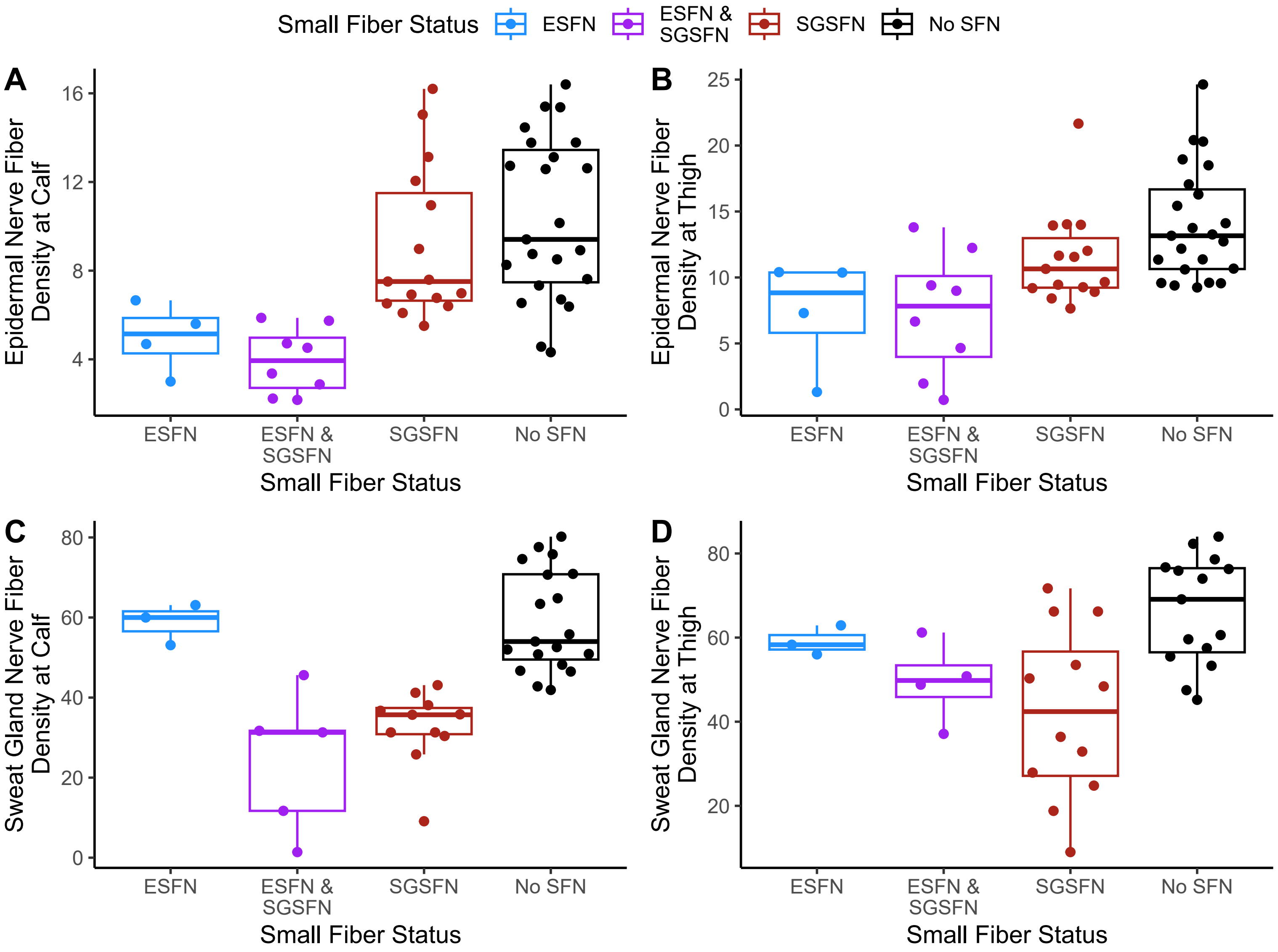


**Supplementary Figure 1 – Skin Punch Biopsy Results by Small Fiber Status.** Epidermal nerve fiber density at the **A)** calf and **B)** thigh by small fiber status, while stratifying dual positive epidermal and sweat gland neuropathy. Sweat gland nerve fiber density at the **C)** calf and **D)** thigh by small fiber status, while stratifying dual positive epidermal and sweat gland neuropathy.

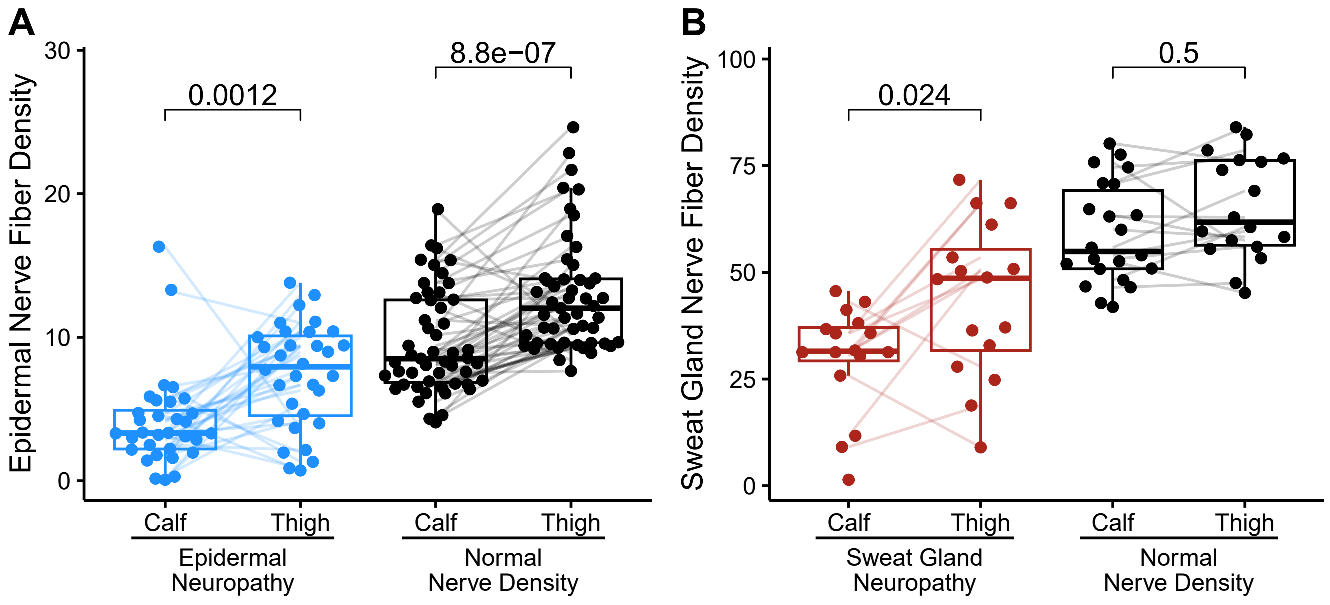


**Supplementary Figure 2 –** Paired measurements of **A)** epidermal nerve fiber density and **B)** sweat gland nerve fiber density at calf and thigh biopsies by neuropathy status. Wilcoxon-signed ranked test used to calculate p-values.

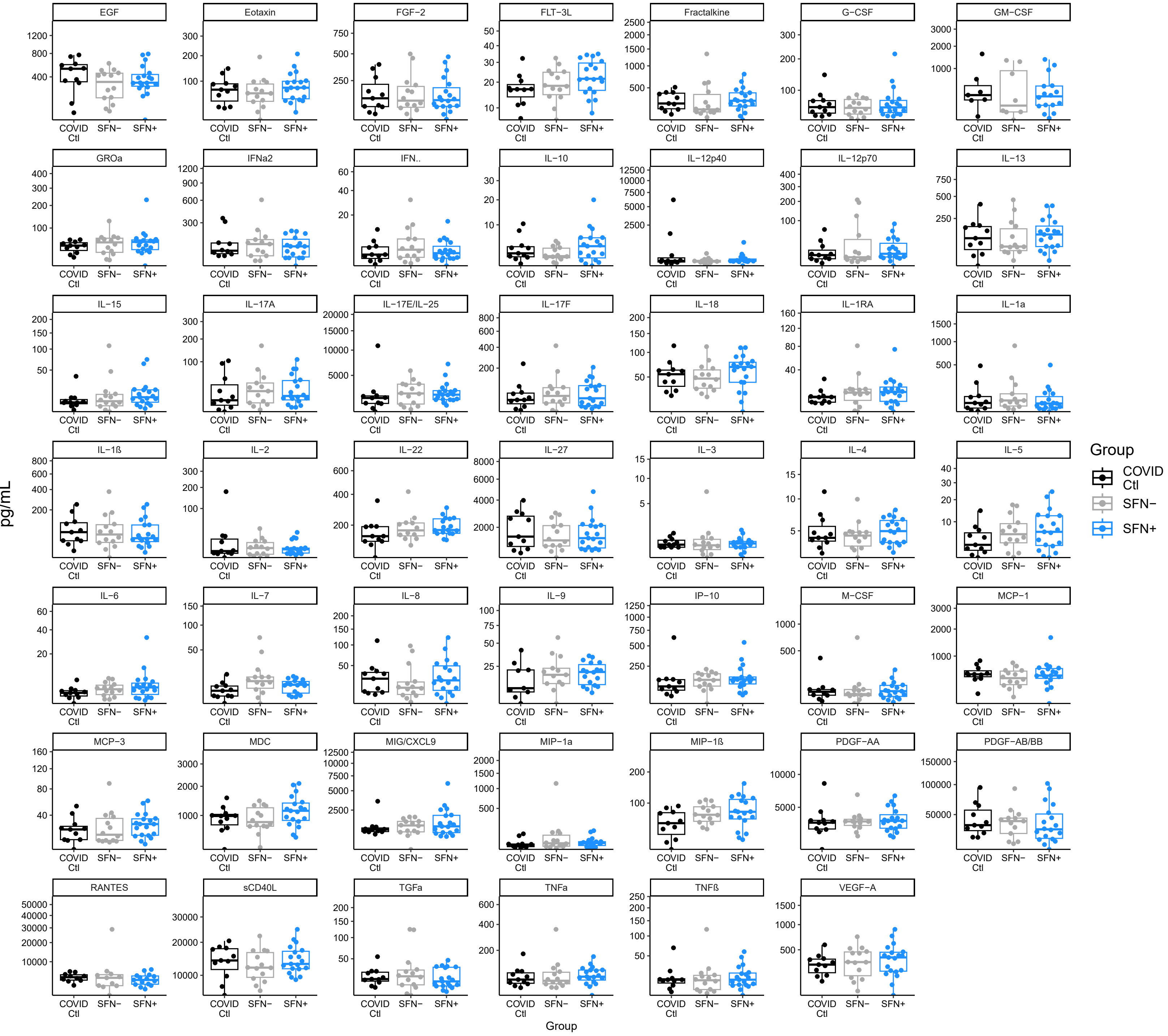


**Supplementary Figure 3 – Cytokine Multiplex Reveals No Difference in 48 Circulating Cytokines in LC Neuropathy Patients.** COVID controls (Ctl) were participants who had a COVID-19 infection, but did not develop LC. There were no significant differences when evaluated with pairwise Wilcoxon rank-sum tests and Benjamini-Hochberg p-value adjustments.

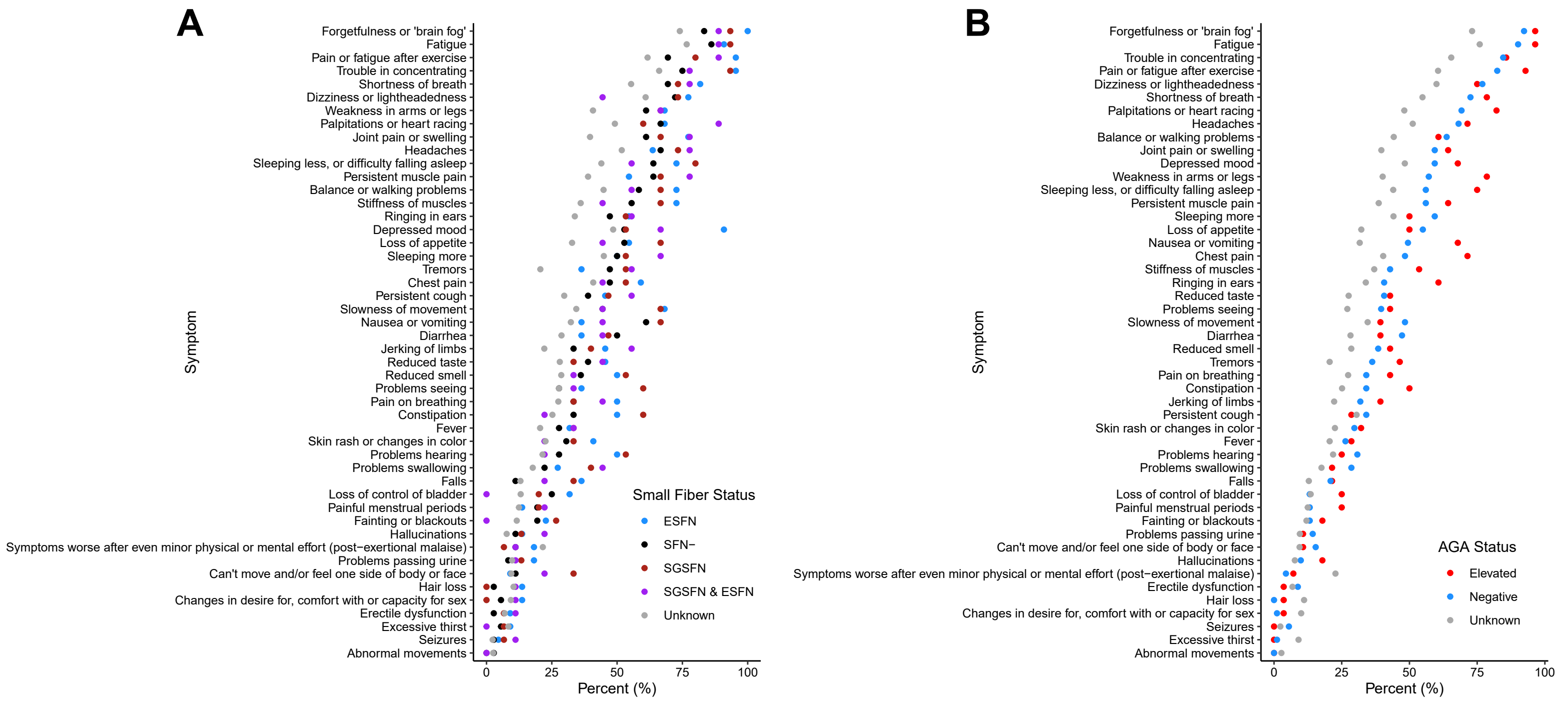


**Supplementary Figure 4 – Other reported LC Symptoms Trend to Differences but Not Significant by AGA Status. A)** Rate of other LC related symptoms by small fiber status. **B)** Rate of other LC related symptoms by AGA positivity. There were no significant differences between the groups (excluding the unknown status group) when evaluated with a chi-square test and Benjamini-Hochberg p-value adjustments.
